# Comparison of Commercially Available and Laboratory Developed Assays for *in vitro* Detection of SARS-CoV-2 in Clinical Laboratories

**DOI:** 10.1101/2020.04.24.20074559

**Authors:** Joshua A. Lieberman, Gregory Pepper, Samia N. Naccache, Meei-Li Huang, Keith R. Jerome, Alexander L. Greninger

## Abstract

Multiple laboratory developed tests and commercially available assays have emerged to meet diagnostic needs related to the SARS-CoV-2 pandemic. To date, there is limited comparison data for these different testing platforms. We compared the analytical performance of a laboratory developed test (LDT) developed in our clinical laboratory based on CDC primer sets and four commercially available, FDA emergency use authorized assays for SARS-CoV-2 (Cepheid, DiaSorin, Hologic Panther, and Roche Cobas) on a total of 169 nasopharyngeal swabs. The LDT and Cepheid Xpert Xpress SARS-CoV-2 assays were the most sensitive assays for SARS-CoV-2 with 100% agreement across specimens. The Hologic Panther Fusion, DiaSorin Simplexa, and Roche Cobas 6800 only failed to detect positive specimens near the limit of detection of our CDC-based LDT assay. All assays were 100% specific, using our CDC-based LDT as the gold standard. Our results provide initial test performance characteristics for SARS-CoV-2 RT-PCR and highlight the importance of having multiple viral detection testing platforms available in a public health emergency.

## Introduction

Since the first infection with SARS-CoV-2 was detected in the United States in January 2020 (1), there has been an exponential growth in cases and deaths (2). At the time of this writing, the US case count exceeds 600,000 with more than 30,000 deaths and considerable geographic heterogeneity (2, 3). Despite social distancing policies, the outbreak of Covid-19, the disease caused by SARS-CoV-2, continues to grow and threatens to overwhelm hospital systems in multiple states (2).

The explosion of Covid-19 cases in the United States has highlighted the critical role diagnostic testing plays in medical and public health decision making in containing and mitigating the SARS-CoV-2 pandemic. Reliable test results enable appropriate utilization of scarce hospital resources, including personal protective equipment (PPE) and negative pressure isolation rooms, as well as public health resources for contact tracing or isolation decision-making (4). In rapid succession in March 2020, multiple assays have become available including both FDA-EUA test platforms and laboratory developed tests (LDTs) for use in high-complexity clinical laboratories. To solve supply chain difficulties, clinical laboratories have had to implement multiple assays using scarce reagent resources, rendering thorough comparisons challenging. A clear understanding of the analytical parameters of these options is important to help guide assay selection by clinical laboratories when supply chain considerations subside (4).

Reverse transcription polymerase chain reaction (RT-PCR) is the mainstay of SARS-CoV-2 detection *in vitro* (5). FDA EUA authorized assays for SARS-CoV-2 have mostly targeted two loci of the positive sense, single-stranded RNA virus by real time RT-PCR and are reported qualitatively. Our laboratory has recently reported that the CDC N2 and WHO E-gene primer/probe sets are among the most sensitive and have detected no false positives in our LDTs (6). FDA EUA authorized platforms use a variety of different primer/probe sets, resulting in the potential for differing analytical sensitivities. In addition to differing analytical sensitivities, commercially available platforms have important operational differences including integrated sample extraction, run time, random access, and acceptable sample types.

Here, the performances of one LDT-EUA assay developed in our clinical laboratory and four FDA-EUA cleared assays were evaluated for detection of SARS-CoV-2. The FDA-EUA cleared assays included were: Hologic Panther Fusion (both RUO and EUA versions, EUA version performed at two study sites), DiaSorin Simplexa COVID-19 Direct (EUA), Cepheid Xpert Xpress SARS-CoV-2 (EUA), and Roche Cobas 6800 (EUA). Test performance characteristics of each RT-PCR were determined compared to our reference LDT assay.

## Materials & Methods

### Specimen collection and consensus panel selection

Nasopharyngeal (NP) swabs (*N* = 169) were collected from patient specimens submitted to the University of Washington Medical Center laboratories for clinical diagnostic testing. LDT performance was validated based on detection of 20 of 20 positives sent by the Washington State Public Health Laboratory in early March. Residual clinical samples were used for validation/verification of each subsequent instrument, including a common panel of 26 specimens (12 positive, 1 inconclusive, and 13 negative) tested at UW by the UW CDC EUA-based LDT (CDC LDT), DiaSorin Simplexa (positives only), Roche Cobas 6800, and tested at LabCorp Seattle on the Cepheid Xpert Xpress, and Panther Fusion (12 positives only). Additional residual (*N* = 115) specimens were tested at UW on individual assays and compared to the reference method (LDT): Panther Fusion (RUO), *N* = 36; Panther Fusion (EUA)-UW, *N* = 20; DiaSorin Simplexa (EUA), *N* = 19; Cobas 6800, *N* = 40. Finally, 28 specimens were used to compare the SARS-CoV-2 assay on the UW Panther Fusion with the DiaSorin Simplexa assay. All same-sample comparisons were performed on specimens stored at 4^°^C for less than 72 hours with no freeze-thaws. Inconclusive results (one of two targets detected) were considered positive due to the high specificity of all assays and limited cross-reactivity seen for SARS-CoV-2 primer sets. This work was approved under a consent waiver from the University of Washington Institutional Review Board.

### Sample processing

For the UW CDC LDT, total nucleic acid (NA) was extracted from 200µL of VTM on the Roche MP96 and eluted in 50µL of elution buffer. Real-time RT-PCR was set up on 5µL of eluate using the CDC N1, N2, and RP (or Exo internal control) primers and run on ABI 7500 real-time PCR instruments as reported previously (6). For the Hologic Panther Fusion, 500µL of VTM was transferred to lysis buffer in manufacturer-provided tubes and loaded directly on the instrument. For the DiaSorin Simplexa and Cepheid Xpert Xpress, 50µL or 300µL of VTM sample respectively was loaded directly into the reaction cartridge with integrated sample process. For the Roche Cobas 6800, 600µL of specimen VTM was added to a barcoded 12×75mm secondary tube and loaded directly on the instrument.

## Results

### Panther Fusion SARS-CoV-2

The Panther Fusion SARS-CoV-2 assay was tested first as research use only (RUO) reagents (*N* = 36), and then again following FDA authorization (*N* = 20). Both Panther Fusion RUO and EUA assays were slightly less sensitive than the CDC-based LDT, missing one positive/inconclusive in each sample set (Table 1a/b). One additional specimen was initially negative with the RUO reagents but was detected upon repeat with the Panther Fusion EUA assay. Discordant specimens were either inconclusive (one target of two detected) or had high average CTs (>37) by the CDC LDT test. All 29 negative specimens generated “Not detected” results by the Hologic Panther Fusion SARS-CoV-2 assay.

**Table 1a.** CDC-based LDT versus Hologic Panther Fusion RUO

| Sample ID | UW LDT |  | Panther Fusion SARS-CoV-2 (RUO) |
| --- | --- | --- | --- |
|  | N1 Ct | N2 Ct | Orf1ab/2ab |
| 56 | 27.2 | 27.8 | 23.2 |
| 07 | 26.2 | 25.5 | 22.8 |
| 46 | 23.9 | 25.8 | 21.4 |
| 81 | 23.8 | 24 | 25.5 |
| 40 | 23.8 | 23.9 | 23.7 |
| 66 | 17.7 | 17.1 | 19.7 |
| 26 | 24.9 | 24.6 | 27 |
| 85 | 35.98 | 35.8 | 33.5 |
| 82 | 29.1 | 29.7 | 29.6 |
| 37 | 23.1 | 22.3 | 22 |
| 70 | 29.7 | 28.9 | 30.5 |
| 29 | 29.4 | 28.2 | 26.3 |
| 68 | 29.4 | 28 | 28.3 |
| 04 | 25.9 | 25.4 | 24.9 |
| 95 | 35 | 37 | 35.3 |
| 55 | 39.2 | 39 | NDET |
| 14 | 36.3 | NDET | 35.5 |

19/19 CDC LDT negatives were negative by Hologic Panther Fusion EUA

**Table 1b.** CDC-based LDT versus Hologic Panther Fusion EUA

| Sample ID | UW LDT |  | Panther Fusion SARS-CoV-2 (EUA) |
| --- | --- | --- | --- |
|  | N1 Ct | N2 Ct | Orf1ab/2ab |
| 100 | 30 | 30.4 | 29 |
| 67 | 20.3 | 20.8 | 19.3 |
| 76 | 27.1 | 27.6 | 31.1 |
| 17 | 23.4 | 23.6 | 20.5 |
| 02 | 18.1 | 17.2 | 19.3 |
| 59 | 29.4 | 29.8 | 31.7 |
| 97 | 18.4 | 17 | 18.9 |
| 52 | 16.8 | 15.9 | 16.9 |
| 90 | 36.1 | 38.4 | NDET/38.1 |
| 79 | 38.4 | NDET | NDET/NDET |

10/10 CDC LDT negatives were negative by Hologic Panther Fusion EUA
NDET, Not Detected

### DiaSorin Simplexa SARS-CoV-2

We next compared the DiaSorin Simplexa SARS-CoV-2 assay to our CDC-based LDT. All 19 specimens (11 positives and 8 negatives) demonstrated complete concordance between the two platforms (Table 2a) with lower CTs recovered by the DiaSorin compared to the LDT on all specimens (average CT difference -2.1 [IQR -2.3 – -1.7]). When we compared SARS-CoV-2 detection on the DiaSorin Simplexa to the Hologic Panther Fusion, all 16 Hologic Panther Fusion positives were detected by the DiaSorin Simplexa, while the DiaSorin Simplexa generated one additional positive in the 12 specimens that were negative by the Hologic Panther Fusion (Table 2b). This discordant specimen was detected by the CDC-based LDT with CTs of 36.8 (N1) and 35.8 (N2), confirming the DiaSorin Simplexa result.

**Table 2a.** CDC-based LDT versus DiaSorin Simplexa EUA

| Sample ID | UW LDT |  | UW DiaSorin |  |
| --- | --- | --- | --- | --- |
|  | N1 Ct | N2 Ct | S-gene | ORF1ab |
| 1A Pos | 27.9 | 27.6 | 25.2 | 25.6 |
| 2A Pos | 18.6 | 19.1 | 18.0 | 18.1 |
| 3A Pos | 27.8 | 27.5 | 25.2 | 26.0 |
| 4A Pos | 28.2 | 27.6 | 25.8 | 26.5 |
| 5A Pos | 31.6 | 31.3 | 28.5 | 29.0 |
| 6A Pos | 33.9 | 34.6 | 31.0 | 31.6 |
| 7A Pos | 31.1 | 31.5 | 28.8 | 29.2 |
| 1C Pos | 34.1 | 33.9 | 31.5 | 32.1 |
| 2C Pos | 33.7 | 34.6 | 32.7 | 32.4 |
| 3C Pos | 32.4 | 32.3 | 29.8 | 30.4 |
| 4C Pos | 34.9 | 34.8 | 32.7 | 33.6 |

8/8 negatives by UW LDT were negative by DiaSorin

**Table 2b.** Hologic Panther Fusion EUA versus DiaSorin Simplex EUA

| Sample ID | Panther Fusion<br>SARS-CoV-2 (EUA) | DiaSorin |  |
| --- | --- | --- | --- |
|  | Orf1ab/2ab | S-gene | ORF1ab |
| 65 | 35.2 | 37.2 | 32.9 |
| 38 | 33.6 | 33.7 | 34 |
| 83 | 31.5 | 28.5 | 27.7 |
| 13 | 30.7 | 27.2 | 27 |
| 39 | 30.1 | 30.7 | 29.2 |
| 10 | 29.6 | 29.1 | 28.1 |
| 56 | 28.7 | 27 | 26.4 |
| 31 | 26.5 | 24.4 | 22.8 |
| 33 | 25.4 | 24.8 | 24.4 |
| 60 | 23.3 | 21.1 | 20.5 |
| 92 | 22.5 | 23 | 23.4 |
| 98 | 21 | 18.1 | 17.3 |
| 40 | 18.2 | 15 | 14 |
| 25 | 17.9 | 16.1 | 15.6 |
| 13 | 16.7 | 15.1 | 14.3 |
| 52 | 15.6 | 13.2 | 12.3 |
| 42* | NDET | 31.8 | 32.7 |

11/12 negatives by Hologic Panther Fusion EUA were negative by DiaSorin

### Roche Cobas SARS-CoV-2

We next compared the Roche Cobas SARS-CoV-2 assay to our CDC LDT. All 20 negatives demonstrated complete concordance between the two platforms (Table 3). One of the 20 positives was not detected by the Roche assay. This specimen had CTs of 38.0 (N1) and 37.4 (N2) in the LDT. Across the 20 positive specimens, CTs were only slightly higher on the Roche Cobas assay compared to the CDC-based LDT, with an average CT difference of 0.6 [IQR -0.1 – 1.5].

**Table 3.** CDC-based LDT versus Cobas 6800 SARS-CoV-2

| Sample ID | UW LDT |  | UW Cobas 6800 |  |
| --- | --- | --- | --- | --- |
|  | N1 Ct | N2 Ct | ORF1ab | E-gene |
| 136 | 26.7 | 27.6 | 26.9 | 27.2 |
| 560 | 16.4 | 16.3 | 19.0 | 19.6 |
| 578 | 23.8 | 24.9 | 25.4 | 26.3 |
| 757 | 23.4 | 24.2 | 24.7 | 25.0 |
| 982 | 20.3 | 20.9 | 21.9 | 22.2 |
| 853 | 20.4 | 21.5 | 21.6 | 21.5 |
| 998 | 14.6 | 15.6 | 15.9 | 16.3 |
| 334 | 20.2 | 21.4 | 21.5 | 21.9 |
| 571 | 18.0 | 18.9 | 17.9 | 18.2 |
| 108 | 35.5 | 35.0 | 31.8 | 34.7 |
| 188 | 36.4 | 35.7 | 35.4 | 37.2 |
| 599 | 24.7 | 25.6 | 26.7 | 27.1 |
| 995 | 25.2 | 26.4 | 26.4 | 26.8 |
| 336 | 29.1 | 29.5 | 31.1 | 31.6 |
| 866 | 31.3 | 31.4 | 31.1 | 32.0 |
| 232 | 36.3 | 36.4 | 32.3 | 35.2 |
| 323 | 28.6 | 28.7 | 29.5 | 30.7 |
| 309 | 14.3 | 15.4 | 14.5 | 14.8 |
| 277 | 19.7 | 21.4 | 20.4 | 20.5 |
| 018 | 38.0 | 37.4 | NDET | NDET |

20/20 negatives by UW LDT were negative by Cobas 6800
NDET, Not Detected

### Five-way same-sample comparison including Cepheid Xpert Xpress SARS-CoV-2 assay

After performing the above pairwise comparisons, we next compared twenty-six specimens (13 positive, 13 negative) from another high-complexity hospital laboratory (LabCorp Seattle). All 26 were also tested on the Cepheid Xpert Xpress SARS-CoV-2 assay (Table 4). All specimens with CT < 35 on the CDC-based LDT were detected by all platforms, and all specimens not detected by the Cepheid Xpert were not detected by two other platforms examined (CDC LDT, Roche Cobas). One of 13 positive specimens was a presumptive positive on the Cepheid assay (E-gene CT 42.6, N2-gene negative); upon repeat per package insert, the N2-gene was detected at CT 42.7 but the E-gene was not detected, yielding a positive result. The CDC LDT demonstrated 100% concordance with the Cepheid Xpert Xpress, also detecting the extremely low viral load specimen above as an inconclusive (N1 37.4, N2 not detected). No other assay detected SARS-CoV-2 RNA in this specimen. In addition, the DiaSorin Simplexa failed to detect a positive specimen that on repeat was detected only by the ORF1ab primer set.

**Table 4.**
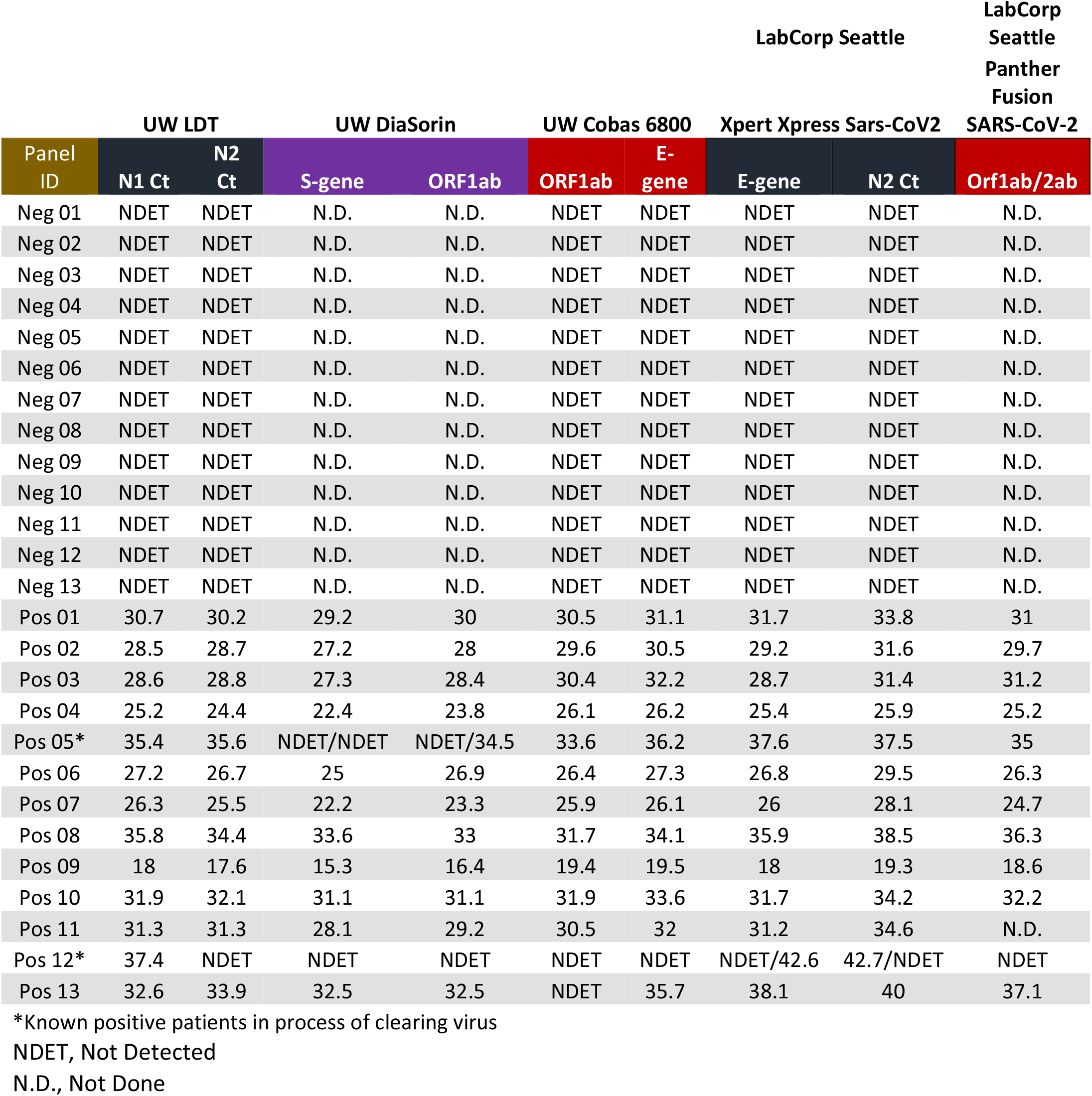
Same-sample comparison of five testing platforms for SARS-CoV-2

## Discussion

This analysis compared the performance characteristics of several *in vitro* diagnostic real time RT-PCR assays to detect SARS-CoV-2 in high-complexity clinical laboratories in one of the early US epicenters of the COVID-19 pandemic. The results demonstrated excellent performance of a CDC-based LDT and the Cepheid Xpert Xpress, concurring with a previous evaluation that demonstrated high sensitivity of the E-gene and N2 primer sets used by the Cepheid assay (6). The Panther Fusion was somewhat less sensitive than either the LDT or the DiaSorin; however, the Panther Fusion detected SARS-CoV-2 RNA in one specimen that was inconclusive (1 of 2 targets detected, thus presumed positive) by the UW CDC LDT. The Roche assay performed on the Cobas 6800 platform detected 28/30 positive samples; both of these discordant specimens had low viral titers (UW CDC LDT *C*_*T*_ >37) and one was the inconclusive specimen. Therefore, we conclude that all the tested assays show good sensitivity for the detection of SARS-CoV-2, with the UW CDC LDT and Cepheid Xpert Xpress SARS-CoV-2 assays having the best and similar sensitivity, followed by the Roche Cobas 6800, DiaSorin Simplexa, and Panther Fusion SARS-CoV-2 assays.

Our results are chiefly limited by the small sample sets used to compare these different assays as well as asynchronous comparisons that only allowed for pairwise comparisons early in the pandemic. For instance, these asynchronous panels most greatly affected our CDC LDT versus Hologic Panther Fusion comparison, which had a greater proportion of high CT positive specimens that resulted in a lower measured sensitivity for the Panther Fusion.. In clinical practice, the minor differences in sensitivity are likely to have little effect on Hologic Panther Fusion SARS-CoV-2 assay performance on VTM specimens, given the CT ranges we have observed in our clinical populations.

Despite their limitations, these data provide a basis for differences in analytical sensitivity at different CTs that may be seen between platforms. For instance, recent reports have demonstrated a slightly higher analytical sensitivity of the Cepheid Xpert Xpress SARS-CoV-2 assay compared to the Roche Cobas SARS-CoV-2 test, and a slightly lower sensitivity of the DiaSorin Simplexa SARS-CoV-2 assay compared to a modified CDC assay, both of which are concordant with our data (7, 8). We also note that, while analytical sensitivity is of critical importance, many other considerations factor into assay platform selection including assay availability, cost, turnaround time, and throughput.

Our results provide an early assessment of performance characteristics of five separate assays for the detection of SARS-CoV-2. During March 2020, reagent availability for SARS-COV-2 RT-PCR assays was heavily constrained, necessitating more limited assay comparisons. All platforms examined here had acceptable performance criteria for testing during the early part of this pandemic. As the supply chain for SARS-CoV-2 RT-PCR attempts to catch up with testing demand, we look forward to additional assay comparison data.

## Data Availability

Available in paper

## References

1. Holshue ML, DeBolt C, Lindquist S, Lofy KH, Wiesman J, Bruce H, Spitters C, Ericson K, Wilkerson S, Tural A, Diaz G, Cohn A, Fox L, Patel A, Gerber SI, Kim L, Tong S, Lu X, Lindstrom S, Pallansch MA, Weldon WC, Biggs HM, Uyeki TM, Pillai SK. 2020. First Case of 2019 Novel Coronavirus in the United States. N Engl J Med 382:929–936.

2. IHME COVID-19 health service utilization forecasting team, Murray CJ. 2020. Forecasting COVID-19 impact on hospital bed-days, ICU-days, ventilator-days and deaths by US state in the next 4 months. preprint, Infectious Diseases (except HIV/AIDS).

3. Dong E, Du H, Gardner L. 2020. An interactive web-based dashboard to track COVID-19 in real time. The Lancet Infectious Diseases S1473309920301201.

4. Babiker A, Myers CW, Hill CE, Guarner J. 2020. SARS-CoV-2 Testing. Am J Clin Pathol.

5. Corman VM, Landt O, Kaiser M, Molenkamp R, Meijer A, Chu DKW, Bleicker T, Brünink S, Schneider J, Schmidt ML, Mulders DGJC, Haagmans BL, van der Veer B, van den Brink S, Wijsman L, Goderski G, Romette J-L, Ellis J, Zambon M, Peiris M, Goossens H, Reusken C, Koopmans MPG, Drosten C. 2020. Detection of 2019 novel coronavirus (2019-nCoV) by real-time RT-PCR. Euro Surveill 25.

6. Nalla AK, Casto AM, Huang M-LW, Perchetti GA, Sampoleo R, Shrestha L, Wei Y, Zhu H, Jerome KR, Greninger AL. 2020. Comparative Performance of SARS-CoV-2 Detection Assays using Seven Different Primer/Probe Sets and One Assay Kit. Journal of Clinical Microbiology.

7. Moran A, Beavis KG, Matushek SM, Ciaglia C, Francois N, Tesic V, Love N. 2020. The Detection of SARS-CoV-2 using the Cepheid Xpert Xpress SARS-CoV-2 and Roche Cobas SARS-CoV-2 Assays. Journal of Clinical Microbiology.

8. Rhoads DD, Cherian SS, Roman K, Stempak LM, Schmotzer CL, Sadri N. 2020. Comparison of Abbott ID Now, DiaSorin Simplexa, and CDC FDA EUA methods for the detection of SARS-CoV-2 from nasopharyngeal and nasal swabs from individuals diagnosed with COVID-19. Journal of Clinical Microbiology.

